# Role of meteorological temperature and relative humidity in the January-February 2020 propagation of 2019-nCoV in Wuhan, China

**DOI:** 10.1101/2020.03.19.20039164

**Authors:** Jose Alvarez-Ramirez, Monica Meraz

## Abstract

Identified in December 2019, the 2019-nCoV emerged in Wuhan, China, and its spread increased rapidly, with cases arising across Mainland China and several other countries. By January 2020, the potential risks imposed by 2019-nCoV in human health and economical activity were promptly highlighted. Considerable efforts have been devoted for understanding the transmission mechanisms aimed to pursue public policies oriented to mitigate the number of infected and deaths. An important question requiring some attention is the role of meteorological variables (e.g., temperature and humidity) in the 2019-nCoV transmission. Correlations between meteorological temperature and relative humidity with the number of daily confirmed cases were explored in this work for the epicenter city of Wuhan, China for the period from 29 January to March 6, 2020. Long-term trend of temperature and relative humidity was obtained with a 14-days adjacent-averaging filter, and lagged correlations of the number of daily confirmed cases were explored. The analysis showed negative correlations between temperatures with the number of daily confirmed cases. Maximum correlations were found for 6-day lagged temperatures, which is likely reflecting the incubation period of the virus. It was postulated that the indoor crowding effect is responsible of the high incidence of 2019-nCoV cases, where low absolute humidity and close human contact facilitate the transport of aerosol droplets.

## 1. Introduction

Starting in December 2019, a group of patients with pneumonia of unknown origin was found in Wuhan, China. The illness was linked to a previously unknown coronavirus, which was named 2019-nCoV (Zhu et al., 2020). In the early days of 2020, the WHO alerted on the potential capacity of the new coronavirus to pose international threatens to human health and global economy. By the end of January 2020, Wuhan, China was positioned as the epicenter of the 2019-nCoV contagion and spreading. In January 31, the number of confirmed cases ascended to about 11950, most of them located in mainland China. Promptly, the coronavirus spread from China to many countries, with about 75 countries reporting confirmed cases. By March 7, the number of confirmed and death cases was about 105,782 and 3,569, respectively.

The availability of the first set of data on the number of detected infected and death cases prompted the early characterization of the propagation dynamics. Zhao et al. (2020) reported the estimated reproduction number R_0_ ranging from 2.24 to 3.58 for the early outbreak phase. Subsequently, Hu et al. (2020) reported the estimated reproduction number 2.68 as of January, 25. Backer et al. (2020) estimated a mean incubation period of 6.4 days. Overall, reports have shown that the 2019-nCoV may have a higher pandemic risk than SARS broken out in 2003. Given the lack of an effective vaccine for controlling the disease, some operational strategies have been envisioned. Tang et al. (2020) used mathematical modeling contrasted to available data (January 29) to suggest that the best measure for reproduction number reduction is persistent and strict self-isolation. However, it has been highlighted that isolation strategies have negative effects in the economic activity, an effect that most governments are reticent to implement, mainly in regions with strong manufacturing activity.

A growing belief, mainly in social networks, is that the 2019-nCoV infection shares some similarities with seasonal flu, and as such the advent of warmer wheatear would weak propagation and fatalities. The motivation behind this folk argument is that influenza epidemic events exhibit wintertime seasonality, with most cases occurring over 2-3 month period between November and March in Northern Hemisphere, and May and September in Southern Hemisphere (Tamerius et al., 2013). Formally, the problem is linked to the role of humidity and temperature in the dynamics of influenza propagation (Lowen and Steel, 2014). Results in this line are still scare, although some reports have pointed out that the transmission of influenza virus is sensible to temperature and humidity (Steel, 2011). Experimental runs on transmission at low (5 °C) versus intermediate (20 °C) temperatures were performed with two influenza B viruses, finding that transmission is more efficient under colder conditions (Pica et al., 2012). It was postulated that transmission of human influenza viruses via respiratory droplet or environment aerosols proceeds most efficiently under cold, dry conditions. Also, the analysis of laboratory and epidemiological data has provided further evidence that temperature plays an important role in the transmission efficiency of influenza viruses (Pica and Bouvier, 2014). Recently, it was found that low humidity and temperature are linked to seasonal influenza activity in the Toronto area (Peci et al., 2019).

Given the potential high risk of the 2019-nCoV, predicting the transmission mechanisms is of prime importance for the timing of implementation of disease prevention and control measures as well as for medical resource allocation (Peci et al., 2019). In this regard, the aim of the present study is to explore the role of environmental factors (temperature and humidity) on the 2019-nCoV activity in Wuhan, china, from 29 January to 6 March, 2020. The present study was motivated by the recent report by Wang et al. (2020), who studied the effect of temperature in the transmission rate of 2019-nCoV and found that every 1 ^°^C increase in the minimum meteorological temperature led to a decrease of the cumulative number of cases by a factor of about 0.86.

## 2. Data sources and methodology

Median temperature and relative humidity (RH) data were obtained from the publicly accessible website www.timeanddate.com/weather/china/wuhan. Data is reported every 6 hours, and mean daily values were obtained by averaging for the four daily values. On the other hand, number of daily new cases and deaths were obtained from the official reports by the Hubei Province Minister of Health at the website http://wjw.hubei.gov.cn. Although official reports are available from 23 January, detailed reports for the Hubei Province are given from 29 January.

The aim of the analysis is to detect co-movements between the number of daily confirmed cases and meteorological variables (temperature and relative humidity). To this end, the estimation of a correlation coefficient will be used to quantify co-movement between two time series. However, since the daily confirmed cases of 2019-nCoV underwent an incubation period (typically 6-8 days), the analysis of co-movement might be biased by lagged effects. In this way, the analysis will be based on the computation of the lagged height cross-correlation analysis (Wang et al., 2016). Briefly, for two time series {*x*_*t*_} and {*y*_*t*_}, *t* = 1,…, *N*, the associated accumulation deviation series are given by

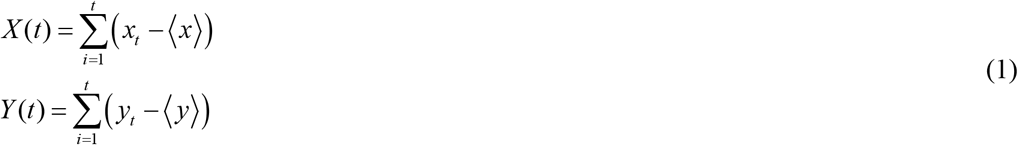

Here, ⟨.⟩ denotes mean values. The cross increment of these time series with interval *L* is computed as

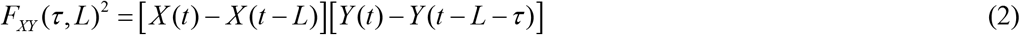

where *τ* denotes the lag. The lagged correlation coefficient is given by

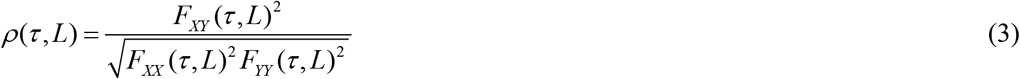

In this way, *ρ* (*τ, L*) ∈[−1,1], with negative values for anti-correlations, and positive values for correlations. It is noted that the analysis is similar to the lagged-DCCA reported by Shen et al. (2015) for two time series. Given the relatively low number of data, the computations should use the scale *L* = *N*. That is, the scrutinized scale corresponds to the number of days from 29 January to 6 March.

## 3. Winter meteorological conditions

Wuhan in mainland China is a metropolis (about 11 millions population), polluted by the intense industrial activity. Climate in Wuhan is temperate, with relatively cold winters. Commonly, cold air can stagnate on the ground. There may be cold periods during which the temperature remains around freezing even during the day, and even snow can fall. Mean min-max temperatures of the winter season are 3-11 ^°^C for December, 1-8 ^°^C for January, 4-11 ^°^C for February and 7-15 ^°^C for March. Figures 1.a and 1.b present respectively the behavior of the temperature and relative humidity for the period from January 1^st^, 2020 (day 1) to date. The mean values of temperature and relative humidity in the period are 7.20±3.85 ^°^C and 78.79±11.68%, respectively. The temperature dynamics exhibits a pattern composed by large oscillations of mean period of about 10 days, and a long-term ascending trend. Similar pattern is presented by the relative humidity dynamics, although the large oscillation has a period of about 15 days. Figure 1.c shows that temperature and relative humidity are weakly negatively correlated (ρ = −0.42).

**Figure 1.**
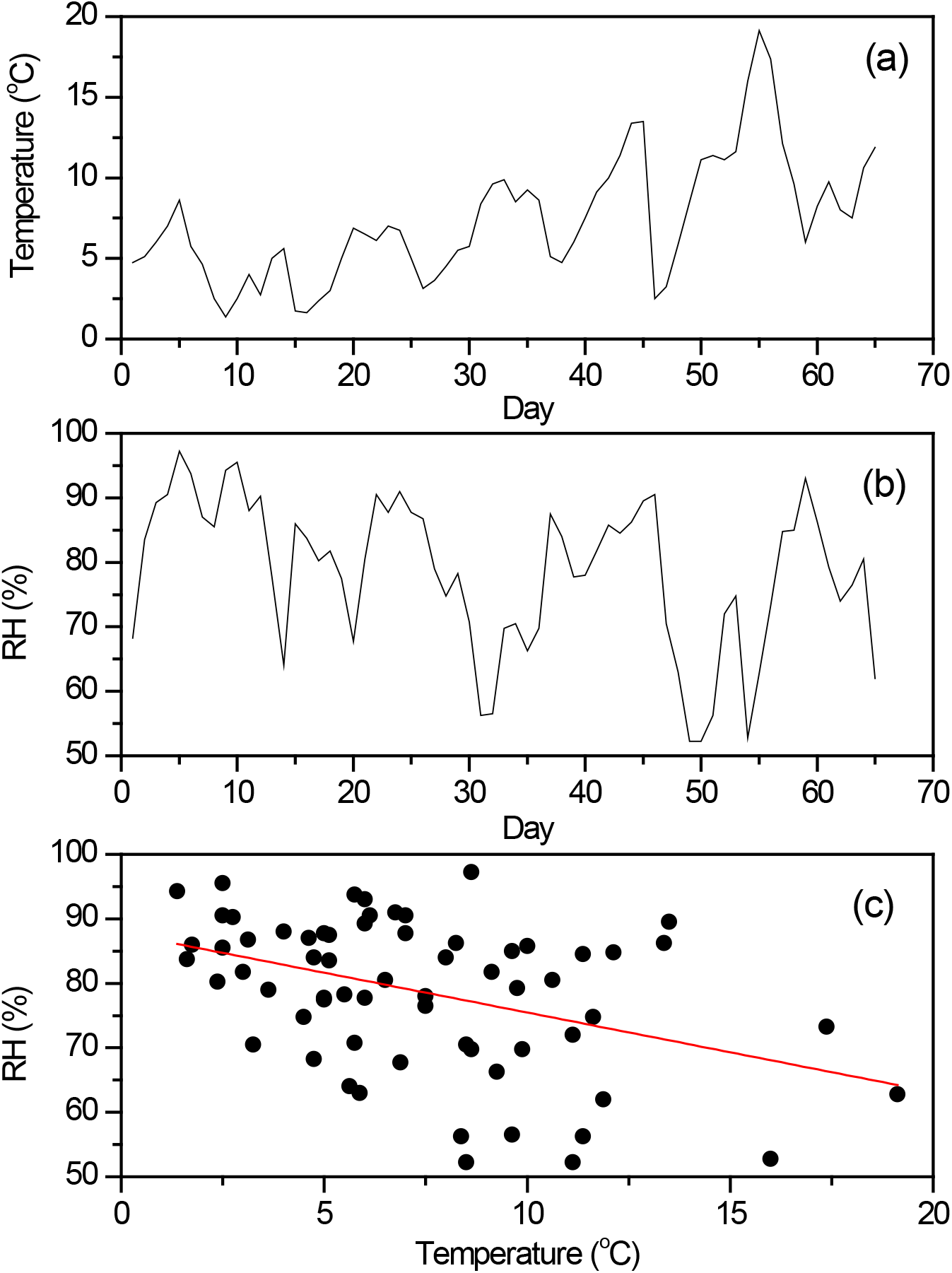
(a) Mean daily meteorological temperature, and (b) relative humidity (RH) in Wuhan, China for the period from 1 January to 6 March, 2020. (c) Plot of the relative humidity versus temperature, showing negative correlations (ρ = −0.42).

## 4. Dynamics of daily confirmed cases and deaths

Figure 2.a shows the dynamics of the daily confirmed cases (C_t_) and deaths (D_t_) for the period from 29 January to 6 March 2020. A large peak in both new cases and deaths at February 12 is exhibited, which corresponds to an improvement of the classification method, when the number of clinically confirmed cases was incorporated to the number of new cases. The new cases showed a positive trend up to February 18, when it presented an important decrease. The behavior of the deaths was similar, although the positive trend was maintained until 23 February. Afterwards, the number of daily deaths has decreased from values of about 100-120 to about 20-30. Figure 2.b shows the plot of the number of deaths (D_t_) and the number of daily confirmed cases (C_t_). The correlations between these two variables is positive (ρ=0.72), a result that can be expected. However, a stronger correlation (ρ=0.85) is exhibited between the number of actual deaths (D_t_) and the number of confirmed cases lagged by 5 days (C_t-5_). The lag of 5 days could be reflecting the mean period between the clinical diagnostic and the death of seriously illness patients. The continuous line in Figure 2.c denotes the least-squares fitting by a quadratic function, where a weak convexity can be observed. This suggests that an increasing number of confirmed cases does not lead linearly to an increasing number of deaths. Figure 2.d shows the correlation coefficient as function of the lag, confirming that maximum linear correlations are exhibited for a lag of about 5 days.

**Figure 2.**
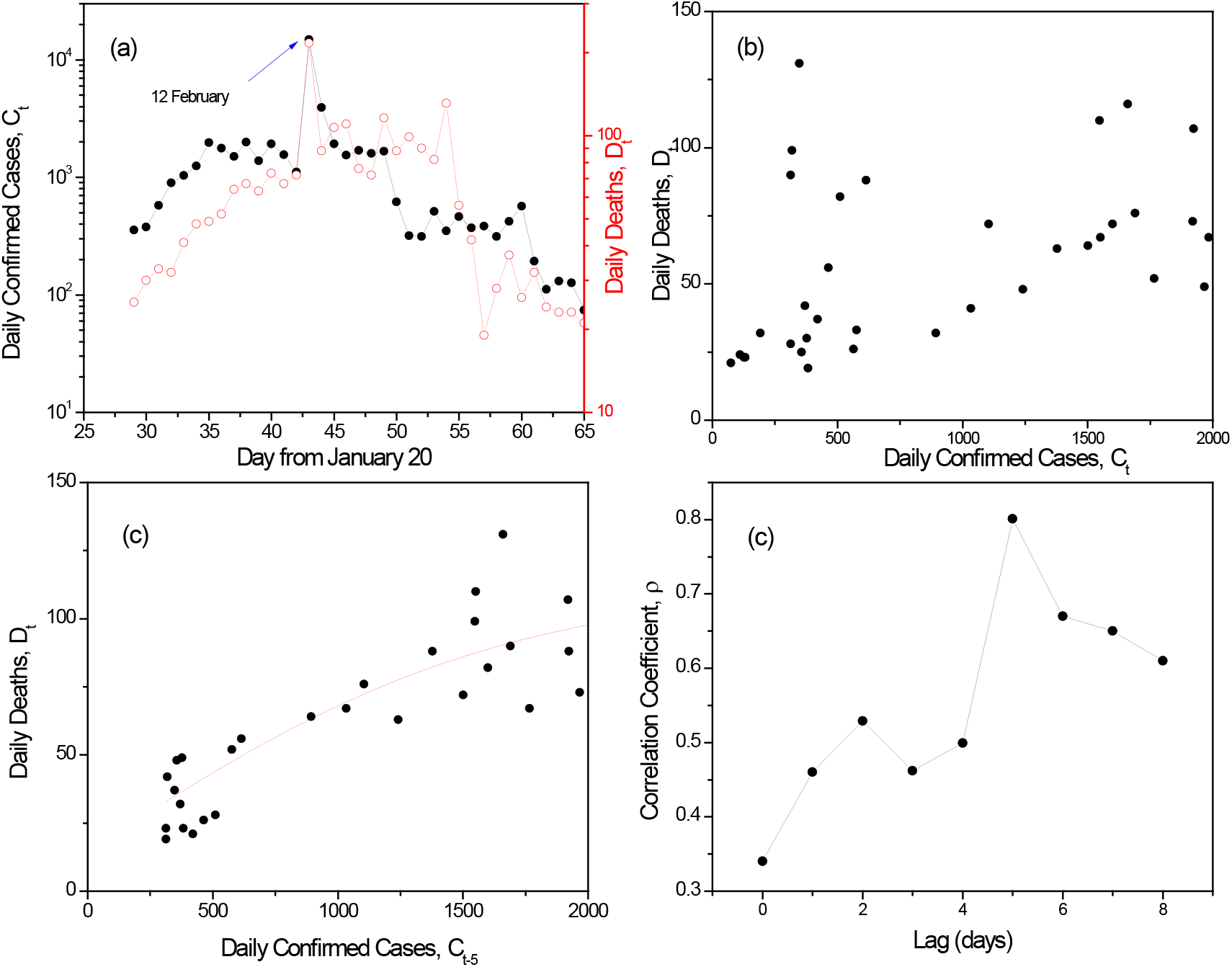
(a) Number of daily confirmed and death cases in Wuhan, China, for the period from 1 January to 6 March, 2020. The large peak in 12 February corresponds to improvements in the detection methods. (b) Plot of contemporaneous daily deaths (D_t_) versus daily confirmed (C_t_) cases. (c) Plot of daily deaths (D_t_) with respect to 5-days lagged daily confirmed cases (C_t-5_). (c) Behavior of the correlation coefficient with respect to the lag in daily confirmed cases.

## 5. Role of temperature and relative humidity

Possible patterns between temperature/relative humidity and the number of new infected cases are explored next. Figure 3.a shows the behavior of the daily confirmed cases (C_t_) with respect to the temperature (T_t_). No discernible regular pattern between the daily confirmed cases and temperature can be observed. The correlation coefficient is very low (ρ = −0.18), reflecting weak negative trend. Similar behavior was exhibited by the new cases with respect to the relative humidity (RH_t_) in Figure 3.b. In this case, ρ = +0.17, indicating the presence of weak positive correlations. Figures 3.c and 3.d present the behavior of the correlation coefficient with respect to the lag for temperature and relative humidity, respectively. Correlations for temperature are negative, with a peak at about 6-7 days. In contrast, correlations for relative humidity are positive, with two prominent peaks at about 3 and 8 days. Nevertheless, the magnitude of the correlations is relative small for both cases. Overall, the results in Figures 3 suggest that no apparent regular pattern is present in the joined dynamics of new infected cases and meteorological variables (temperature and relative humidity). In the face of this feature, a different strategy was pursued. A key observation is that the meteorological variables showed large fluctuations along a secular trend (see Figure 1). Today confirmed cases correspond to infected cases in past days. It has been reported that the incubation period of COV-19 is typically 7-14 days, and could be as long as 27 days. In this regard, the effects of temperature in today confirmed cases are not pointwise, but distributed along several past days. That is, the dynamical behavior of today confirmed cases is likely to be affected by the temperature trend in the past few days. The proposed strategy consists in considering the secular trend of the temperature and relative humidity dynamics, and to search patterns with respect to lagged values.

**Figure 3.**
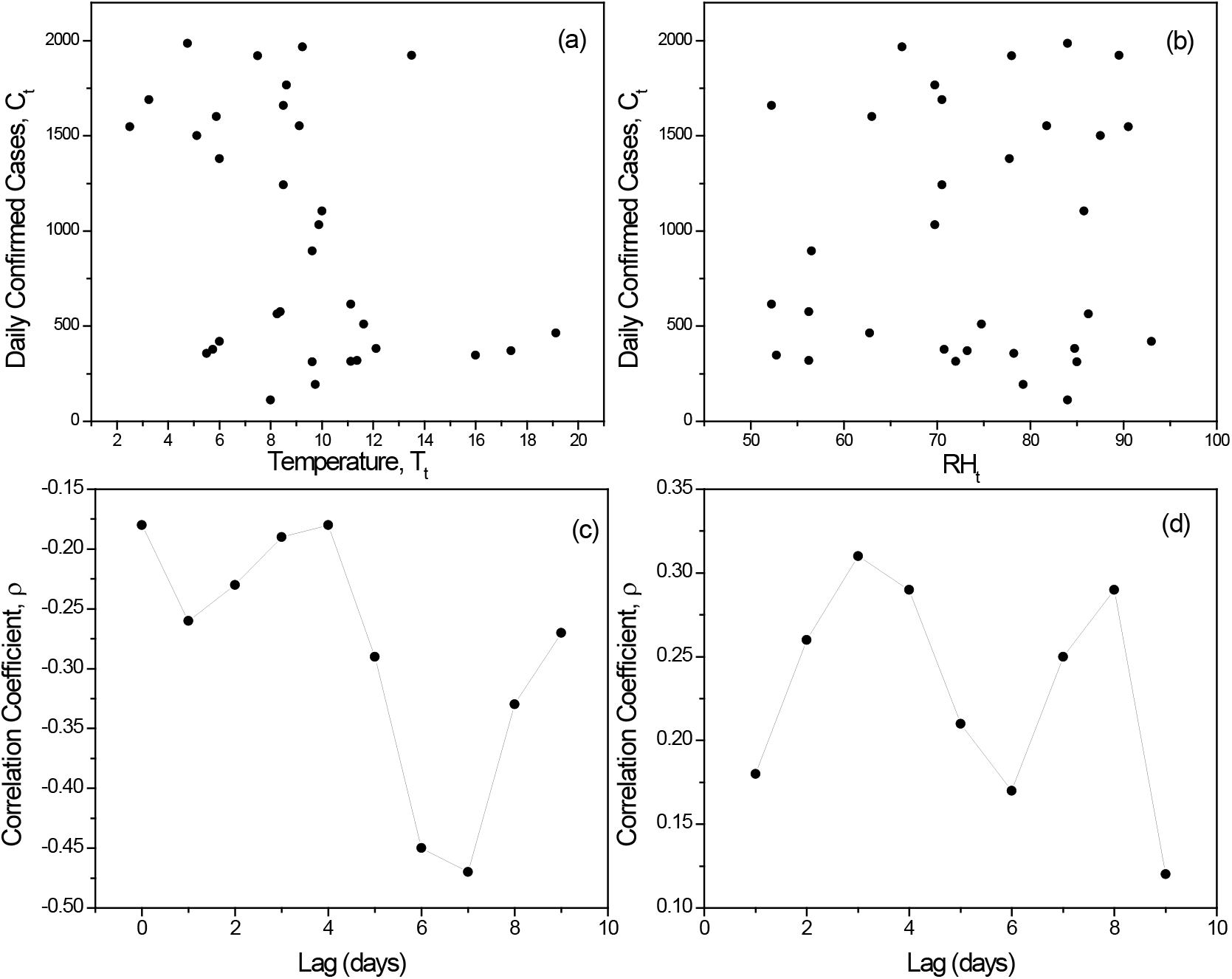
Plots of daily confirmed cases versus (a) temperature and (a) relative humidity. The plots exhibit a scattered pattern with weak correlations. The behavior of the correlation coefficient with respect to the lag for (c) temperature and (d) relative humidity exhibit a peak in magnitude for lags of about 5-8 days.

Figures 4.a and 4.b present respectively the long-term trend of temperature and relative humidity, which were obtained by means of a 14^th^-order moving-average filtering. From January 10, the temperature showed a positive trend, until February 28 when the trend was negative. The trend temperature goes from about 4-5 ^°^C by January 10, to about 10-11 ^°^C by February 24. In contrast, the relative humidity showed a negative trend, until February 24 when the trend was positive. Figure 4.c compares the trends of temperature and relative humidity, showing an apparent negative correlation (ρ = 0.81) between these two signals. That is, the increase of temperature was accompanied by a decrease of the relative humidity.

**Figure 4.**
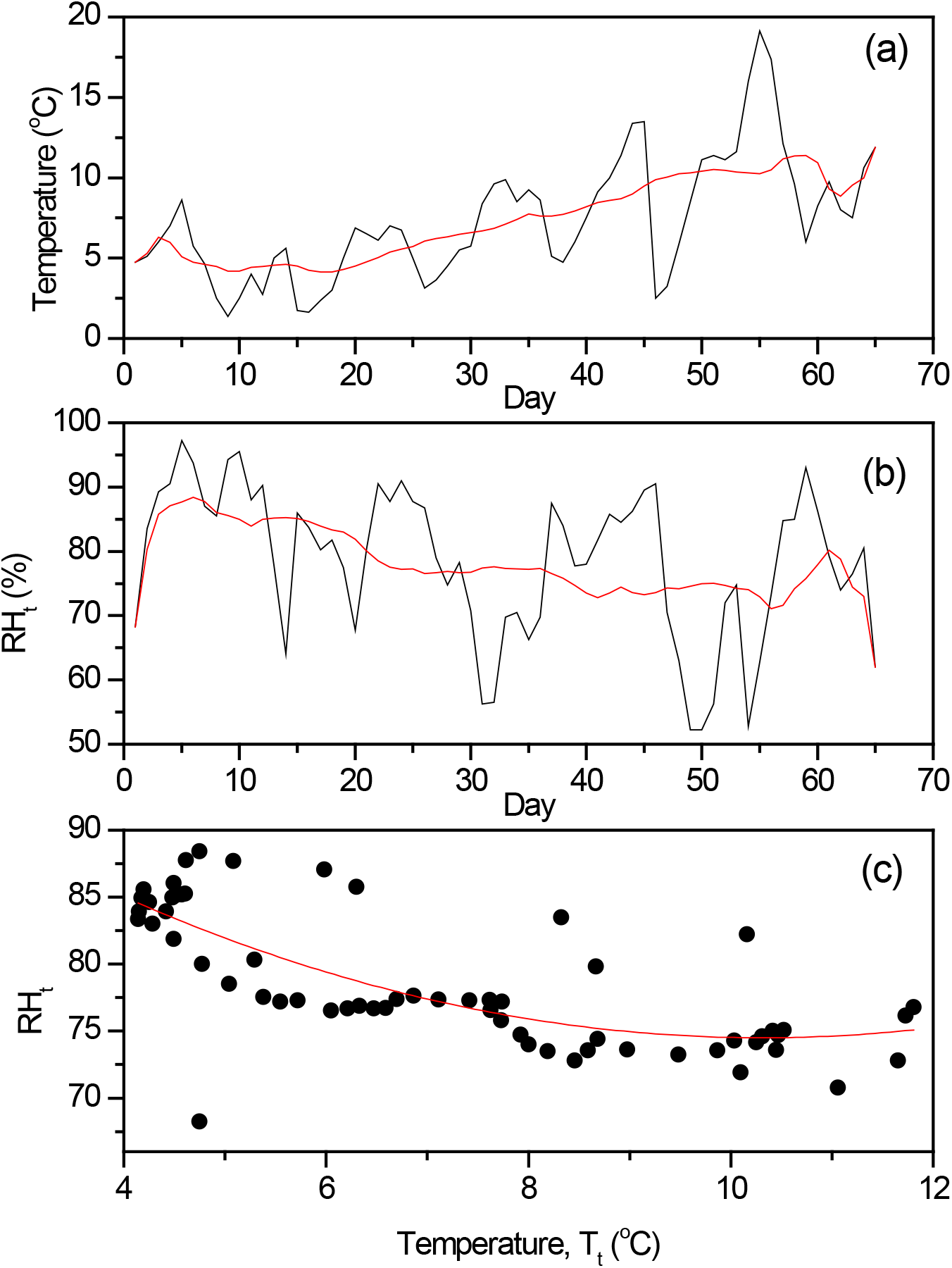
Meteorological (a) temperature, and (b) relative humidity in Wuhan, China, for the period from 1 January to March 6, 2020. The red lined depict the long-term trend obtained by 15-days moving average filtering. (c) Plot of the trend relative humidity and the trend temperature. A negative correlation between these trends can be observed.

Figures 5.a and 5.b present the behavior of the correlation coefficient with respect to the lag for temperature and relative humidity, respectively. A pattern is more discernible than in figures 3.c and 3.d, indicating that the use of temperature trend instead of daily temperature is more appropriate for the analysis of correlations between daily confirmed cases and meteorological variables. The correlation coefficient for temperature is negative and its magnitude achieved a maximum (ρ = −0.77) at about 6 days. The correlation coefficient for relative humidity is positive and S-shaped, achieving the maximum value (ρ = + 0.65) at about 6-7 days. For the lag where maximum correlations were detected, figures 5.c and 5.d show the plot of the number of daily confirmed cases versus trend temperature and relative humidity, respectively. This result indicates that the today number of daily confirmed cases is correlated with the temperature and relative humidity lagged about one week. The lag of 6-7 days could correspond to the mean incubation period (Tian et al., 2020). The pattern displayed by temperature in Figure 5.c is interesting. The continuous line depicts the least-squares sigmoid fitting, which shows a sharp transition from high to low number of daily confirmed cases. Relative to the 6-days lagged trend temperature, the number of daily confirmed cases shows two phases with crossover temperature of about 8.6 ^°^C. Below this temperature, the mean number of daily confirmed cases is about 1635 per day, and above such temperature value the number of daily confirmed cases decreased to about 360 per day. Overall, the results described in Figure 5 are in line with a recent report by Wang et al. (2020), who found that the 1 ^°^C increase in the minimum meteorological temperature led to a decrease of the cumulative number of cases by 0.86 in 429 studied cities.

**Figure 5.**
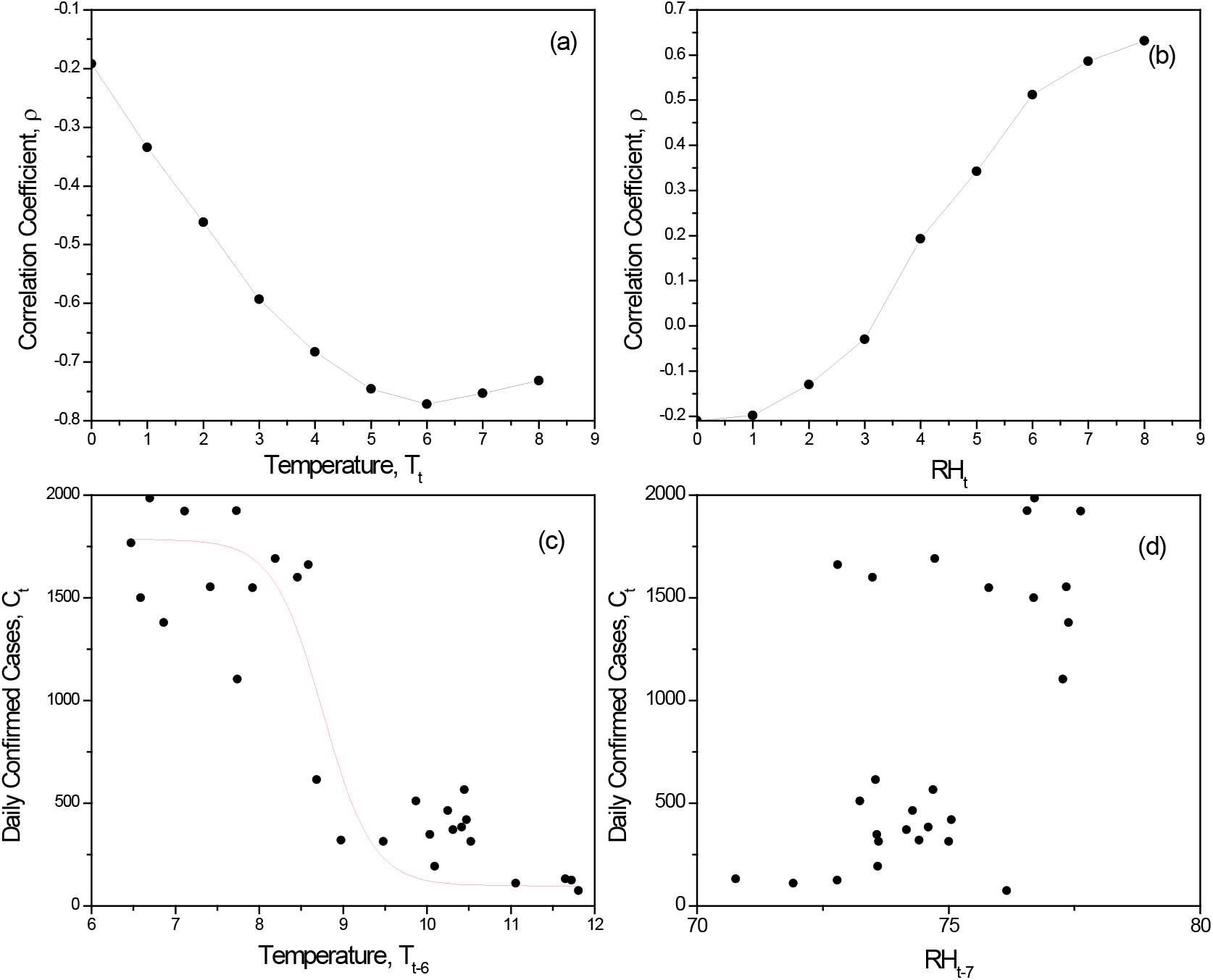
Behavior of the correlation coefficient with respect to the lag, for (a) trend temperature and (b) trend relative humidity. (c) Plot of daily confirmed cases and 6-day lagged trend temperature. (d) Plot of daily confirmed cases and 7-day lagged trend relative humidity.

## 6. Discussion

Figure 5.c showed the presence of a marked pattern between the daily confirmed cases and 6-days lagged temperature trend. In turn, this shows that environmental temperature plays an important play in the transmission dynamics of 2019-nCoV in Wuhan, China. The 6-7 days period in the temperature lag is likely reflecting the mean incubation period of the virus (Backer et al., 2020). On the other hand, the two-phase pattern of the number of daily confirmed cases showed that a large incidence of daily confirmed cases (about 1650 per day) occurred for low environment temperatures (lower than about 8 ^°^C). Sajadi et al. (2020) reported that 2019-nCoV spread is persistent in cities and regions along a 30-50^°^ N’ corridor showing consistent meteorological conditions with average temperatures of 5-11 ^°^C, combined with low specific (3-6 g/kg) and absolute humidity (4-7 g/m^3^). Also, Wang et al. (2020) reported that 1 ^°^C increase in temperature and 1% increase in relative humidity lower the effective reproduction number by 0.0383 and 0.0224, respectively.

A sharp decrease of the number of daily confirmed cases (from about 1800 to about 350 cases per day) was observed for higher temperatures. Oliveiros et al. (2020) found that the doubling time of newly confirmed cases has positive correlation with temperature and negative with humidity. In turn, this suggests a decrease in the rate of transmission of 2019-nCoV with spring and summer arrivals in the North Hemisphere. The important reduction of the number of new cases is not necessarily indicating that the virus stability is sensible to relatively high temperatures, but that the human transmission network is affected by the environment temperature. It is likely that the sharp transition in Figure 5.c is reflecting the so-called indoor crowding effect. Average low outdoor temperatures result commonly in indoor crowding as people activities takes place primordially in, e.g., homes, commercial malls, sport events, and cinema, where temperature is regulated to temperate conditions (about 10-25 ^°^C). Although the outdoor relative humidity is high in winter, the indoor absolute humidity hardly changes after artificial calefaction mechanisms. As a consequence, the indoor ambient is warm and dry, which are benign conditions for virus transmission under respiratory droplet or aerosol route mechanisms (Schaffer et al., 1976; Soebiyanto et al., 2015). Warm and dry indoor conditions affect positively the transport of respiratory droplets. In fact, stable respiratory droplets under low absolute humidity and warm temperature has typical diameter < 5 μm, which remain airborne for prolonged periods (Tellier, 2009). Besides, small aerosol droplets within the microns range moves randomly, increasing the distance and exposure time over which transmission can occur. In this way, Figure 5.c suggest that the high transmission rate of 2019-nCoV in Wuhan at low temperatures (less than about 8 ^°^C), China could be explained from an increase of indoor human activity, which is realized under regulated ambient conditions characterized by warm temperature and low absolute humidity, which facilitate virus transmission. The relatively high dispersion of the data points in Figure 5.c for low temperature might be linked to confounding factors, including precipitation, air pollution and improvements in medical facilities. However, a major effect of the meteorological temperature cannot be ruled out. Whether or not the results found in this study can be extended to other cities is an issue that should be addressed in order to pursue public policies to mitigate and contend with the virus spreading. For instance, mathematical modeling has suggested that persistent and strict self-isolation is recommended for cutting off transmission channels (Tang et al., 2020). Under the hypothesis of indoor crowding effect, a further recommendation involves the development of improved calefaction and air conditioning equipment for controlling both temperature and absolute humidity. The use of traditional calefaction and air conditioning devices could be accompanied by humidity controls to avoid absolute and relative humidity depletion, and in this way reduce the risk of virus transmission. However, the task temperature and humidity ranges are still unclear. Bu et al. (2020) studied SARS data and arrived to the conclusion that persistent warm and dry weather is conducive to the survival of the 2019-nCoV and postulated that temperature ranging 13-19 °C and humidity ranging 50-80% are suitable conditions for the survival and transmission of coronavirus.

## 7. Conclusions

Predicting the transmission dynamics of 2019-nCoV is of prime importance for a proper design of public health policies intended for disease prevention and control measures as well as for economical and medical resource allocation. The present study found insights that the environmental temperature plays an important role in the transmission rate of the 2019-nCoV. It was postulated that the indoor crowding effect is the main responsible of the high transmission rate of 2019-nCoV in Wuhan, China in the period January-February 2020.

## Data Availability

Data used in the manuscrito are available under request to authors.

